# Reduction of lymphocyte at early stage elevates severity and death risk of COVID-19 patients: a hospital-based case-cohort study

**DOI:** 10.1101/2020.04.02.20050955

**Authors:** Jun Fei, Lin Fu, Ying Li, Hui-Xian Xiang, Ying Xiang, Meng-Die Li, Fang-Fang Liu, De-Xiang Xu, Hui Zhao

**Affiliations:** Second Affiliated Hospital, Anhui Medical University, Hefei, Anhui Province, 230032, China; Department of Toxicology, Anhui Medical University, Hefei, Anhui Province, 230032, China; Union Hospital, Tongji Medical College, Huazhong University of Science and Technology, Wuhan, Hubei Province, 430040, China

**Keywords:** Coronavirus disease 2019, Severe acute respiratory syndrome coronavirus-2, Multiple organ injuries, Severity, Death risk

## Abstract

**Background and objective:** Severe acute respiratory syndrome coronavirus-2 (SARS-CoV-2)-induced coronavirus disease 2019 (COVID-19) has been pandemic worldwide. Several reports observed a reduction of lymphocytes among COVID-19 patients. However, clinical significance of lymphocyte reduction in COVID-19 patients remains unclear. The objective of this study was to analyze the association between lymphocyte reduction at early stage and the prognosis of COVID-19 patients.

**Methods:** All 192 hospitalized patients with COVID-19 were enrolled. Electronic medical records, including demographic data, clinical characteristics, comorbidities and exposure history, were collected. Biochemical indexes on admission and chest computed tomography (CT) were detected. Patient’s prognosis was followed up.

**Results:** On admission, 84 (43.8%) patients suffered from lymphocyte reduction among COVID-19 patients. The count and percentage of lymphocytes on admission were lower among more than seventy-year-old patients than those of younger patients. Multivariate logistic regression revealed that older age was a risk factor of lymphocyte reduction. Of interest, chest CT score, a key marker of lung injury, was increased among COVID-19 patients with lymphocyte reduction. By contrast, PaCO2, SpO2 and oxygenation index, several respiratory function markers, were decreased in COVID-19 patients with lymphocyte reduction. Moreover, TBIL and DBIL, two markers of hepatic injury, creatinine and urea nitrogen, two indices of renal function, and creatine kinase, AST and LDH, three myocardial enzymes, were elevated in COVID-19 patients with lymphocyte reduction. Among 84 COVID-19 patients with lymphocyte reduction, 32.1% died. Fatality rate was obviously higher in COVID-19 patients with lymphocyte reduction than those with normal lymphocyte (*RR*=5.789, *P*<0.001).

**Conclusion:** Older COVID-19 patients are more susceptible to lymphocyte reduction. Lymphocyte reduction at early stage aggravates the severity of multiple organ injuries and elevates death risk of COVID-19 patients.

---

In December 2019, a cluster of cases of pneumonia emerged in Wuhan City in Central China’s Hubei Province. Genetic sequencing of isolates obtained from patients with pneumonia identified a novel coronavirus as the etiology, now officially classified as severe acute respiratory syndrome coronavirus 2 (SARS-CoV-2) *(1)*. Subsequently, the new disease has been officially named as corona virus disease 2019 (COVID-19) by WHO *(2-4)*. Since COVID-19 broke out in Wuhan City, SARS-CoV-2 injection has been pandemic in the whole world. As of March 18, 2020, total 81151 confirmed cases have been identified and 3242 patients died from SARS-CoV-2 infection in China. Moreover, 114408 confirmed cases and 4651 death cases were found in other countries *(5)*.

The clinical symptoms and features of COVID-19 patients include fever, dry cough, bilateral lung ground glass opacity, dyspnea and diarrhea. In severe and critical ill cases, SARS-CoV-2-caused pneumonia leads to not only severe acute respiratory syndrome but also multiple organ failure even death *(6)*. Furthermore, several studies found that lymphocyte count from peripheral blood was decreased in most patients with COVID-19. Lymphocyte count in severe patients was obviously lower than that of common patients *(1, 6, 7)*. Nevertheless, what factors lead to lymphopenia remains unclear. The association between lymphocyte reduction and the prognosis of COVID-19 patients needs to be determined.

The objective of this study was to analyze influence factors of lymphocyte reduction as well as the association between lymphocyte reduction and prognosis of COVID-19 patients in a hospital-based case-cohort study. Our results showed that older age was a risk factor of lymphocyte reduction on admission. We found that lymphocyte reduction on admission aggravated the severity of multiple organ injuries and elevated death risk of COVID-19 patients. This research provides evidences for the first time that peripheral lymphocyte count on admission may be a potential indicator of prognosis for COVID-19 patients.

## Methods

### Subjects

In the present research, 220 cases confirmed with SARS-CoV-2 infection were enrolled from Union Hospital of Huazhong University of Science and Technology from January 1 to January 30, 2020. Union Hospital of Huazhong University of Science and Technology is one of COVID-19-designated hospitals for the hospitalization of probable and confirmed COVID-19 cases in Wuhan City in central China’s Hubei Province. Medical team from the Second Affiliated Hospital of Anhui Medical University zealously went to Union Hospital and taken part in medical treatment of COVID-19 cases in Wuhan City. We excluded cases with a negative SARS-CoV-2 RNA detection result, incomplete data or testing result. Children and pregnant women were also excluded from this research. At last, 192 patients involved in this trial and 28 cases eliminated. We reviewed and collected clinical electronic medical records including demographic data, clinical characteristics, comorbidities, nursing records and disease exposure history for all patients with laboratory confirmed SARS-CoV-2 infection. Biochemical indexes on admission and chest computed tomography (CT) were detected. We followed up the outcomes of COVID-19 patients. This study was approved by the institutional ethics board of the Second Affiliated Hospital of Anhui Medical University and Union Hospital of Huazhong University of Science and Technology. All participants or guardians gave written informed consent.

### Date collection

Demographic data, laboratorial results and CT image findings were acquired from medical records in the Union Hospital of Huazhong University of Science and Technology. The demographic data of all patients were collected: gender, age, smoking history, exposure history, comorbidities including cardiovascular disease, hypertension, chronic pulmonary disease, hepatic disease, diabetes and other chronic diseases, symptoms and signs including: cough, fever, fatigue and diarrhea. Biochemical indexes on admission included: blood routine, oxygen saturation, partial pressure of carbon dioxide (PaCO2) in artery, partial pressure of oxygen (PaO2), fraction of inspiration O□ (FiO2), PaO2/FiO2 ratio (oxygenation index), total bilirubin (TBIL), direct bilirubin (DBIL), alanine aminotransferase (ALT), total protein, albumin, globulin, albumin/globulin ratio, creatinine, urea nitrogen, uric acid, aspartate aminotransferase (AST), AST/ALT ratio, creatine kinase, lactate dehydrogenase, cardiac troponin □, D-dimer, erythrocyte sedimentation rate (ESR), C-reactive protein (CRP). All patients underwent chest CT scan. The total CT images were analyzed by two independent radiologists with rich clinical experience. All images were viewed on both lung (width, 1500 HU; level, −700 HU) and mediastinal (width, 350 HU; level, 40 HU) settings. The presence or absence of all image features was recorded: ground-glass opacities, consolidation, traction bronchiectasis, bronchial wall thickening, reticulation, subpleural bands, vascular enlargement and lesion distribution. The detailed CT image characteristics of 200 COVID-19 patients were described in other work *(8)*. On the image scans, the pulmonary tissues were divided into two single lungs (left and right lung) and three zones (upper, middle, and lower zones of lung in the bilateral lungs). These areas of the lung were defined as the “upper zones” above the level of the carina; the “middle zones” between the level of the carina and the level of the inferior pulmonary veins; and the “lower zones” below the level of the inferior pulmonary veins. The CT score was determined by visually estimating the extent of disease in each zone. The severity scores were evaluated according to the percentage of lung parenchyma in each zone that showed evidence of each recorded abnormality: (1) no injury; (2) 1 point, involvement of less than 25% of the image; (3) 2 ponits, 25% to 50%; (4) 3 points, 50% to 75%; (5) 4 ponits, more than 75%. A total severity scores (between 0 and 24) for each lung were generated via adding all the partial areas. The total scores represented the damage area for each lung tissue *(9)*.

### Statistical analysis

All data were statistically analyzed with SPSS 21.0 software. Categorical variables were expressed with frequencies and percentages. Data were expressed as mean or median for the continuous variables. Means for continuous variables were compared with independent-samples *t* tests when the data were normally distributed; if not, the Mann-Whitney test was used. The comparison of discrete variables between different groups was evaluated using the *chi-square* and *Fisher’s* exact test. Moreover, the main risks related with lymphocyte reduction were examined using multivariable logistic regression model. Statistical significance was determined at *p* values of less than 0.05.

## Results

### 1. Demographics data and clinical characteristics among COVID-19 patients

All the 192 patients (49.5% males) aged between 22 to 87 years were included in the analysis. The normal range of lymphocyte count in the peripheral blood is from 0.8 to 4 (10^9^/L). The demographic and clinical characteristics of all COVID-19 patients are showed in Table 1. Patients with high and low counts of lymphocyte (split at the low thresholds, 08×10^9^/L) were on difference in gender, comorbidities including smoker, diabetes, hypertension, hepatic disease, cardiac disease, pulmonary disease and other chronic disease. Clinical symptoms, such as fever, diarrhea, fatigue and cough, as well as injured pulmonary nodes (bilateral lungs, single left lung, single right lung) were no difference in COVID-19 patients with normal lymphocyte and lymphocyte reduction (Table 1). Nevertheless, the number of patients under 49 years old with lymphocyte reduction was less than those with normal lymphocyte [11(13.1%) vs 37(34.3%)]. On the contrary, the number of patients over 70 years old with lymphocyte reduction was more than those with normal lymphocyte [24(28.6%) vs 14(16.0%)].

**Table 1.** Demographics and baseline characteristics of patients with COVID-19.

| Characteristic | Lymphocyte reduction<br>(N=84) | Normal lymphocyte<br>(N=106) | *P* |
| --- | --- | --- | --- |
| Gender (N, %) |  |  | 0.056 |
| Male | 49 (58.3) | 48 (44.4) |  |
| Female | 35 (41.7) | 60 (55.6) | 0.304 |
| Smoking (N, %) |  |  | 0.023 |
| Yes | 29 (34.5) | 54 (50.9) |  |
| No | 55 (65.5) | 52 (49.1) |  |
| Age (N, %) |  |  | 0.001 |
| < 59 year | 37 (44.0) | 59 (55.7) |  |
| 60-69 year | 23 (27.4) | 33 (31.1) |  |
| > 70 year | 24 (28.6) | 14 (13.2) |  |
| Comorbidities (N, %) | 72 (84.7) | 83 (77.6) | 0.153 |
| Diabetes | 55 (65.5) | 74 (68.5) | 0.525 |
| Hypertension | 40 (47.6) | 56 (51.9) | 0.476 |
| Hepatic disease | 5 (6.0) | 4 (3.7) | 0.735 |
| Cardiac disease | 10 (11.9) | 6 (5.6) | 0.114 |
| Pulmonary disease | 6 (7.1) | 2 (1.9) | 0.073 |
| Other disease | 3 (3.6) | 4 (3.7) | 0.941 |
| Symptoms (N, %) |  |  |  |
| Fever | 70 (83.0) | 95 (89.6) | 0.203 |
| Diarrhea | 48 (57.1) | 65 (60.7) | 0.560 |
| Fatigue | 46 (54.8) | 56 (51.9) | 0.688 |
| Cough | 39 (46.4) | 52. (48.1) | 0.537 |
| Chest CT abnormal (N, %) |  |  |  |
| Bilateral lungs | 83 (98.8) | 105 (99.1) | 0.868 |
| Left lung | 80 (95.2) | 99 (93.3) | 0.589 |
| Right lung | 81 (96.4) | 103(97.2) | 0.772 |

### 2. Older COVID-19 patients are more susceptible to lymphocyte reduction

The blood routine of the whole COVID-19 patients on admission to hospital were measured and the number of abnormal cases was calculated. As shown in Table 2, we found that 84 (43.8%) cases’ lymphocyte count were decreased, 97 (50.8%) cases’ lymphocyte percentage were decreased and 4 (2.1%) were increased. 64 (33.5%) cases’ white blood cell (WBC) were under the normal range and 15 (7.9%) above the normal range. 105 (55.2%) cases’ eosinophils count and 120 (62.5%) eosinophils percentage were below the normal range. 114 (59.4%) neutrophil percentage were higher than the upper limit. The effects of different gender on lymphocyte count and lymphocyte percentage were analyzed. As shown in Table 3, lymphocyte counts were similar in male and female patients with COVID-19. Lymphocyte percentage in males were obviously lower than those in females (16.1% vs 22.3%, *P*=0.012). The effects of ages on lymphocyte count and lymphocyte percentage were then evaluated. The lymphocyte counts in patients over 70 years old were evidently lower compared with those under 60 years old and from 60 to 69 years old (0.66 vs 0.92 with 0.92, *P*=0.012). Similarly, the lymphocyte percentage in patients over 70 years old were lower than those under 60 years old and between 60 to 69 years old (12.1% vs 22.1% with 15.8%, *P*=0.001). Besides, there were no differences of lymphocyte count and lymphocyte percentage in the smokers and non-smokers. Among all cases, 163 (81.5%) had at least one of comorbidities, such as 137 (68.5 %) diabetes, 101 (51.5%) hypertension, 9 (4.5%) hepatic disease, 16 (8.0%) cardiac disease, 8 (4.0%) pulmonary disease and 7 (3.5%) other chronic diseases. The impacts of comorbidities on lymphocyte count and lymphocyte percentage were then analyzed. There were no statistical difference of lymphocyte count and lymphocyte percentage between COVID-19 patients with and without diabetes, hypertension, hepatic disease and other chronic diseases (Table 3). Interestingly, we found that lymphocyte percentage in patients with cardiac disease and pulmonary disease were prominently lower than those in patients without cardiac disease and pulmonary disease (12.0% vs 20.6%, *P*=0.028; 10.6% vs 20.1%, *P*=0.045). Multivariable logistic regression was used to analyze risk factors of lymphocyte reduction in 192 patients with COVID-19. As shown in Table 4, the *OR* of lymphocyte reduction in patients from 60 to 69 years old was 0.290 (95% *Cl*: 0.096, 0.874; *P*=0.028), the *OR* of lymphocyte reduction in patients under 70 years old was 0.350 (95% *Cl*: 0.124, 0989; *P*=0.048). No significant correlation was observed between gender, smoker with comorbidities and lymphocyte reduction in COVID-19 patients (Table 4).

**Table 2.** Blood routine on admission to hospital of COVID-19 patients.

| Parameter | Normal range | Outside the range (N, %) |  |
| --- | --- | --- | --- |
|  |  | Below | Above |
| WBC ($10^9/L$) | 4-10 | 64 (33.5) | 15 (7.9) |
| Neutrophil ($10^9/L$) | 2-7 | 30 (15.7) | 23 (12.0) |
| Lymphocyte ($10^9/L$) | 0.8-4 | 84 (43.8) | 0 |
| Monocytes ($10^9/L$) | 0.12-1.2 | 4 (2.1) | 2 (1.0) |
| Eosinophil ($10^9/L$) | 0.02-0.5 | 105 (55.2) | 1 (0.5) |
| Basophil ($10^9/L$) | 0-0.1 | 0 | 5 (2.6) |
| Neutrophil (%) | 50.0-70.0 | 9 (4.7) | 114 (59.4) |
| Lymphocyte (%) | 20.0-40.0 | 97 (50.8) | 4 (2.1) |
| Monocytes (%) | 3.0-12.0 | 10 (5.2) | 10 (5.2) |
| Eosinophil (%) | 0.5-5.0 | 120 (62.5) | 0 |
| Basophil (%) | 0-1.0 | 4 (2.1) | 0 |

**Table 3.**
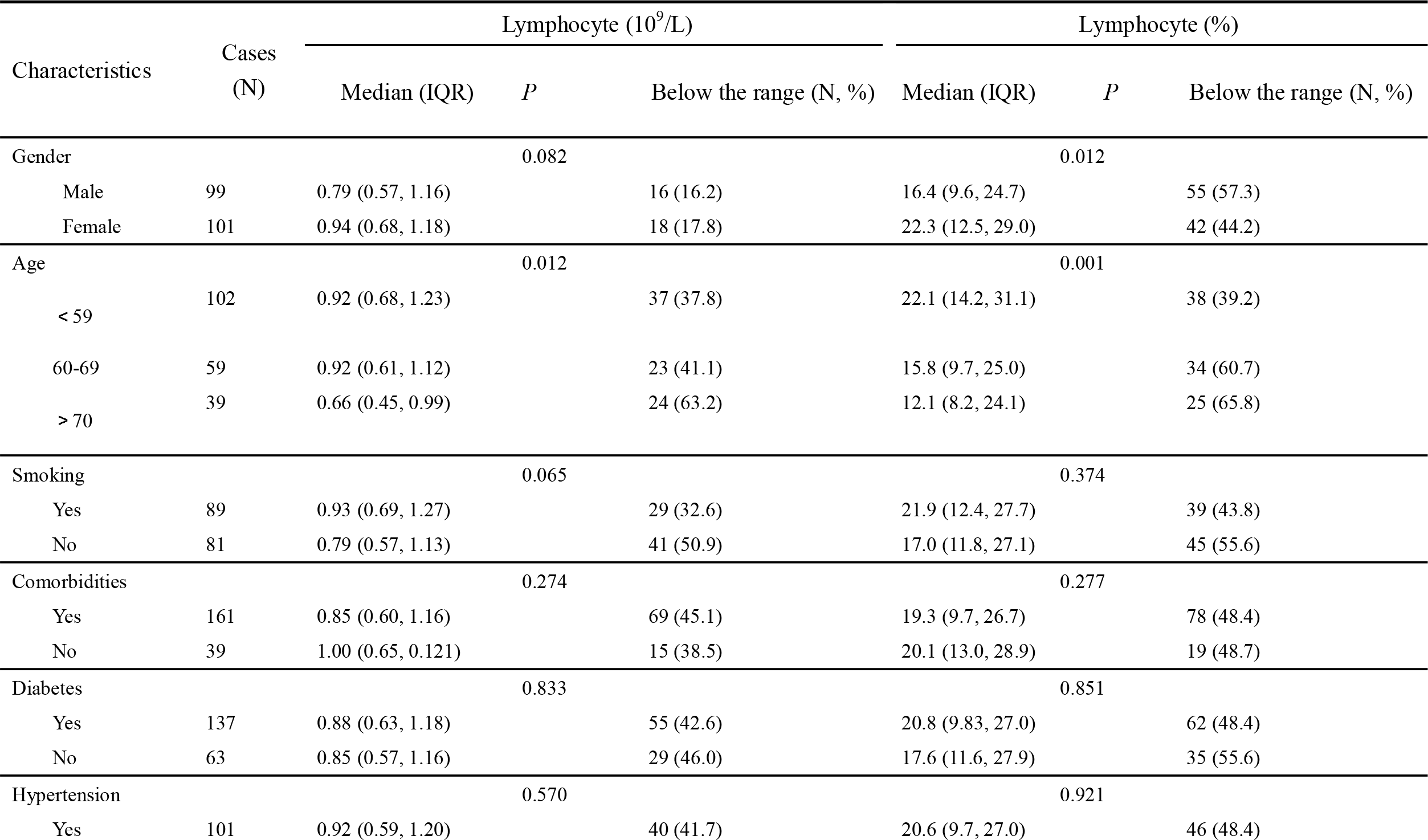
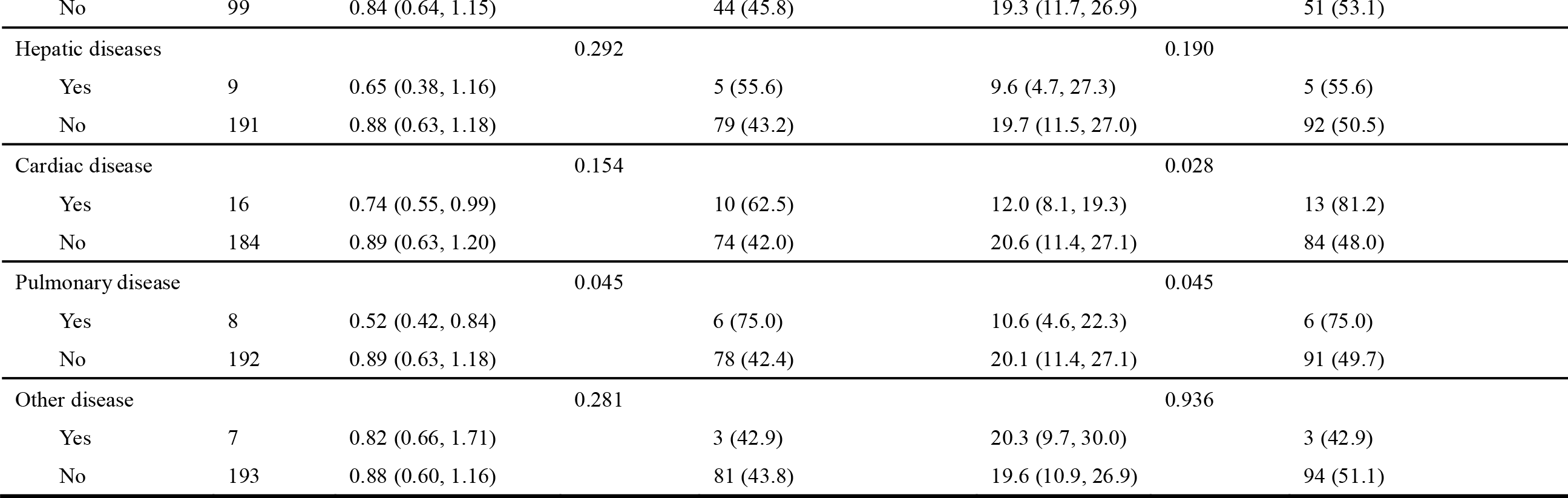
The count and percentage of lymphocyte on admission among COVID-19 patients.

**Table 4.** Multivariable logistic regression between demographics data and lymphocyte reduction on admission in COVID-19 patients.

| Parameter | $\beta$ | Wald | *P* | *OR* (95% CI) |
| --- | --- | --- | --- | --- |
| Gender (Male) | 0.498 | 1.428 | 0.232 | 1.645 (0.727, 3.720) |
| Smoking | -0.479 | 0.979 | 0.322 | 0.619 (0.240, 1.600) |
| Age |  |  |  |  |
| 60-69 | -1.239 | 4.837 | 0.028 | 0.290 (0.096, 0.874) |
| > 70 | -1.049 | 3.922 | 0.048 | 0.350 (0.124, 0.989) |
| Comorbidities |  |  |  |  |
| Diabetes | 0.072 | 0.024 | 0.878 | 1.075 (0.427, 2.707) |
| Hypertension | -0.104 | 0.065 | 0.798 | 0.901 (0.406, 2.002) |
| Hepatic diseases | 0.462 | 0.267 | 0.605 | 1.587 (0.276, 9.145) |
| Cardiac disease | 0.597 | 0.637 | 0.425 | 1.817 (0.419, 7.884) |
| Pulmonary disease | 0.972 | 1.123 | 0.289 | 2.642 (0.438, 15.932) |
| Other disease | 0.647 | 0.521 | 0.470 | 1.910 (0.330, 11.076) |

### 3. Lymphocyte reduction on admission aggravates lung damage among COVID-19 patients

Chest CT were examined among COVID-19 patients. As shown in Figure 1A, mild lung markings and patch clouding lung ground glass shadow in the subpleural area of bilateral lungs were found in COVID-19 patients with normal lymphocyte. Whereas, increased pulmonary brochovascular shadows, indistinct lung markings and blur, pulmonary consolidation and nodules, and extensive high-density area with obvious lung ground glass shadow in the bilateral lungs were the main lung manifestations in COVID-19 patients with lymphocyte reduction (Figure 1B). Besides, the severity of lung damage was scored by two experienced radiologists blind to the clinical data. As shown in Figure 1C-1E, the results showed that left lung CT score, right lung CT score and bilateral lungs CT score were all evidently increased in patients with lymphocyte reduction (5.0 vs 3.0, 6.0 vs 3.0, 11.0 vs 6.0). Further correlation analysis indicated that lymphocyte count was significantly and negatively associated with CT scores of left lung, right lung and bilateral lung (r=−0.341, *P*< 0.001; r=−0.396, *P*<0.001; r=−0.435, *P*<0.001), respectively.

**Figure 1.**
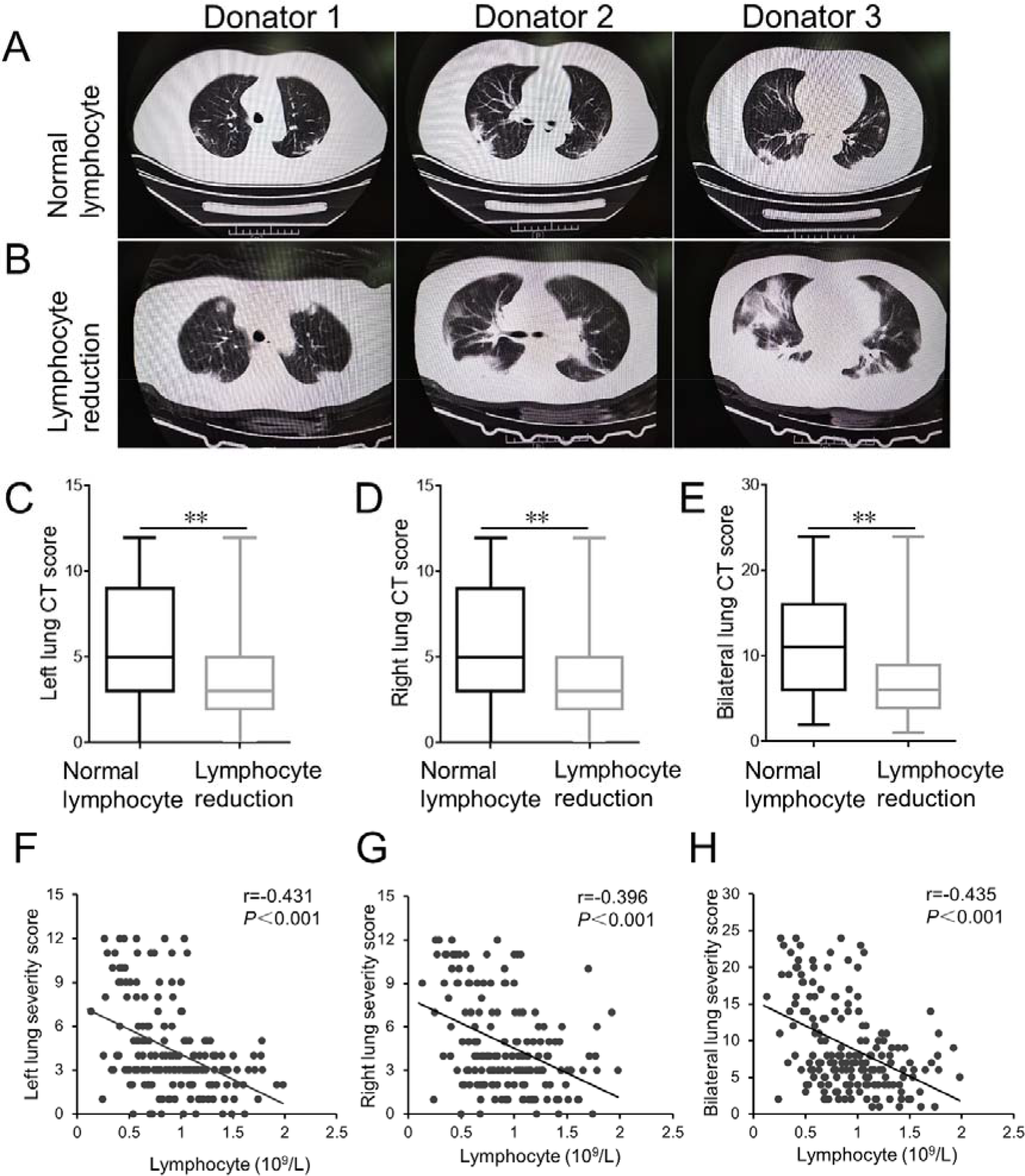
The CT images and the association between lymphocyte reduction and CT scores. (A) CT images from three different patients with normal lymphocyte were showed. (B) CT images from three different patients with lymphocyte reduction were showed. (C-E) CT scores of left lung, right lung and bilateral lung were evaluated. (F-H) Associations between lymphocyte count and CT scores in left lung, right lung and bilateral lung were analyzed among COVID-19 patients (N=192). \*\**P*<0.01.

### 4. Lymphocyte reduction aggravates extrapulmonary multiple organ injuries among COVID-19 patients

The effect of lymphocyte reduction on admission on extrapulmonary organ multiple organ injures was analyzed in COVID-19 patients. Hepatic function related biochemical indexes on admission were detected among all COVID-19 patients. As shown in Table 5, TBIL and DBIL in patients with lymphocyte reduction were obviously higher than those in patients with normal lymphocyte. The level of ALT was no difference between two groups. Compared with patients with lymphocyte reduction, total protein and albumin were lower than those in patients with normal lymphocyte. There was no difference in globulin and albumin/globulin ratio between two groups. Renal functions were also measured on admission among COVID-19 patients. As shown in Table 5, creatinine and urea nitrogen were increased in patients with lymphocyte reduction. No significant difference on uric acid was observed between two groups. Moreover, myocardial function results suggested that creatine kinase, LDH, AST and AST/ALT ratio were significantly increased in patients with lymphocyte reduction compared with in patients with normal lymphocyte. The numbers of myoglobin-positive and cardiac troponin □-positive patients with lymphocyte reduction were more than the patients with normal lymphocyte. The number of creatine kinase isoenzymes-positive patients were equal between two groups. Besides, respiratory function related biochemical indexes of COVID-19 patients were evaluated on admission. PaCO2, SpO2, oxygenation index were remarkably decreased in patients with lymphocyte reduction. In addition, we found that D-dimer and erythrocyte sedimentation rate were prominently increased in patients with lymphocyte reduction.

**Table 5.** Biochemical indexes on admission to hospital of COVID-19 patients.

| Parameter | Lymphocyte reduction<br>(N=84) | Normal lymphocyte<br>(N=108) | *P* |
| --- | --- | --- | --- |
| **Hepatic function markers** |  |  |  |
| TBIL (μm/L) | 12.9 (9.6, 18.5) | 10.8 (8.8, 15.0) | 0.006 |
| DBIL (μm/L) | 4.2 (2.7, 6.1) | 2.9 (2.1, 4.0) | < 0.001 |
| ALT (U/L) | 23.5 (16.0, 49.3) | 22.0 (15.0, 41.0) | 0.373 |
| Total protein (g/L) | 60.4 (56.9, 65.0) | 62.5 (59.4, 68.4) | < 0.001 |
| Albumin (g/L) | 34.8 (32.5, 38.0) | 37.8 (33.9, 41.9) | 0.001 |
| Globulin (g/L) | 26.2 (22.0, 28.9) | 26.7 (23.3, 29.1) | 0.327 |
| Aibumin/globulin ratio | 1.3 (1.1, 1.7) | 1.5 (1.2, 1.8) | 0.134 |
| **Renal function markers** |  |  |  |
| Creatinine (μm/L) | 78.0 (62.0, 98.0) | 63.0 (52.0, 77.0) | < 0.001 |
| Urea nitrogen (mm/L) | 5.7 (4.1, 7.5) | 3.9 (3.1, 5.5) | < 0.001 |
| Uric acid (μm/L) | 257.0 (189.0, 338.0) | 257.0 (190.0, 318) | 0.762 |
| **Myocardial function markers** |  |  |  |
| Creatine kinase (U/L) | 107.0 (59.3, 217.5) | 79.0 (49.0, 138.0) | 0.031 |
| Creatine kinase isoenzymes (N, %) | 57 (80.3) | 86 (86.9) | 0.063 |
| Myoglobin (N, %) | 43 (64.2) | 72 (74.2) | 0.030 |
| Cardiac troponin □ (N, %) | 50 (79.4) | 80 (87.9) | 0.032 |
| LDH (U/L) | 347.0 (254.0, 483.0) | 231.0 (173.0, 311.3) | < 0.001 |
| AST (U/L) | 40.5 (26.8, 68.3) | 31.0 (22.0, 46) | 0.001 |
| AST/ALT ratio | 1.6 (1.3, 2.2) | 1.3 (1.0, 1.8) | 0.002 |
| **Respiratory function markers** |  |  |  |
| PaCO<sub>2</sub> (mmgH) | 33.5 (30.0, 39.0 ) | 39.0 (36.0, 43.0) | < 0.001 |
| SpO<sub>2</sub> (%) | 74.0 (54.3, 102.5) | 94.0 (71.0, 158.0) | 0.002 |
| Oxygenation index | 237.7 (162.8, 326.2) | 338.1 (238.6, 426.6) | < 0.001 |
| PH value | 7.44 (7.41, 7.47) | 7.44 (7.41, 7.47) | 0.526 |
| **Other parameters** |  |  |  |
| D-dimer (ng/mL) | 661.5 (411.3, 3154.3) | 459.5 (330.8, 962.8) | 0.003 |
| ESR (mm/h) | 49.0 (28.0, 77.0) | 33.5 (20.3, 58.0) | 0.003 |
| CRP (mg/L) | 46.2 (15.4, 95.2) | 53.6 (10.5, 89.2) | 0.497 |

### 5. Lymphocyte reduction on admission elevates disease severity of COVID-19 patients

The effect of lymphocyte reduction on admission on disease severity of COVID-19 was analyzed. Among all patients, severe and critical ill cases, defined as oxygenation index lower than 300, accounted for 107 (55.9%). Common cases, oxygenation index higher than 300, accounted for 85 (44.1%). The relationship between lymphocyte count and disease severity of COVID-19 was analyzed. As shown in Table 6, the severe or critical ill patients were 59 (72.0%) in patients with lymphocyte reduction, and were 43 (41.0%) in patients with normal lymphocyte. The *RR* was 1.757 (95% *Cl*: 1.346, 2.292; *P*<0.001) in severe and critical ill COVID-19 cases with lymphocyte reduction.

**Table 6.** The relationship between lymphocyte reduction and the severity with death risk in COVID-19 patients.

| Characteristics | Total cases, N | Severe/Critical ill patients, N (%) | *RR (95% CI)* | *P* |
| --- | --- | --- | --- | --- |
| Lymphocyte reduction | 84 | 59 (72.0) | 1.757 (1.346, 2.292) | < 0.001 |
| Normal lymphocyte | 108 | 43 (41.0) | 1 | — |
| Characteristics | Total cases, N | Dead patients, N (%) | *RR (95% CI)* | *P* |
| Lymphocyte reduction | 84 | 27 (32.1) | 5.789 (2.504, 13.367) | < 0.001 |
| Normal lymphocyte | 108 | 6 (5.56) | 1 | — |

### 6. Lymphocyte reduction on admission elevates the death risk of COVID-19 patients

The effect of lymphocyte reduction on admission on the fatality rate was evaluated. As shown in Table 6, 32.1% (27/84) died in COVID-19 patients with lymphocyte reduction, whereas only 5.56% (6/108) died in patients with normal lymphocyte. Further analysis showed that lymphocyte reduction elevated death risk of COVID-19 patients. The *RR* was 5.789 (95% *Cl*: 2.504, 13.367; *P*<0.001) in COVID-19 patients with lymphocyte reduction.

## Discussion

The present research analyzed the association between lymphocyte reduction at early stage and prognosis of COVID-19 patients in a hospital-based case-cohort study. The main findings of the present research include: older COVID-19 patients are more susceptible to lymphocyte reduction; lymphocyte reduction on admission aggravates the severity of multiple organ injuries among COVID-19 patients; lymphocyte reduction on admission elevates death risk of COVID-19 patients.

Several studies found that lymphocyte count was decreased in COVID-19 patients (1, 7, *10, 11)*. In addition, several reports showed that lymphocyte count was lower in severe COVID-19 patients than in common ones *(12)*. What factors lead to lymphocyte reduction on admission among COVID-19 patients remains obscure. In the present research, we analyzed the relationship between age and lymphocyte reduction among COVID-19 patients. Our results showed that almost half of patients’ lymphocyte count and percentage were reduced to below normal range. Moreover, the number and percentage of lymphocytes were lower in COVID-19 patients over 70 years old than those of younger patients. The effects of gender and comorbidities on lymphocyte reduction were analyzed among COVID-19 patients. Our results showed that lymphocyte percentage were obviously decreased in males than those of females. Of interest, lymphocyte percentage was reduced only in COVID-19 patients with either cardiac diseases or pulmonary chronic diseases. To control possible confounding factors, multivariable logistic regression was used to further analyze the influence of gender, age and comorbidities on lymphocyte reduction in COVID-19 patients. We found that older age was the major risk factor of lymphocyte reduction on admission among COVID-19 patients. Our results suggest that lymphocytes may be a direct target of SARS-CoV-2. Elderly patients are more susceptible to suffer from lymphocyte reduction at the early stage of SARS-CoV-2 infection.

Previous studies found that multiple organ injures were prevalent in almost all COVID-19 patients, especially in critical ill cases *(13-15)*. In the present research, we evaluated the severity of lung injury in COVID-19 patients using CT scores. We observed an obvious lung damage in almost all COVID-19 patients. Moreover, the CT scores were higher in COVID-19 patients with lymphocyte reduction than those in COVID-19 patients with normal lymphocyte. Correlation analysis revealed that lymphocyte counts were negatively associated with CT scores. The association between lymphocyte reduction and respiratory function indices was then analyzed among COVID-19 patients. As expected, PaCO2, SpO2 and oxygenation index, several respiratory function markers, were decreased in COVID-19 patients with lymphocyte reduction. Next, the association between lymphocyte reduction and extrapulmonary organ injuries was analyzed among COVID-19 patients. We found that TBIL and DBIL, two markers of hepatic injury, creatinine and urea nitrogen, two indices of renal function, and creatine kinase, AST and LDH, three myocardial enzymes, were obviously elevated in COVID-19 patients with lymphocyte reduction. These results suggest that lymphocyte reduction at the early stage may be a potential indicator of multiple organ injures among COVID-19 patients.

Several reports demonstrated that lymphocyte reduction was more obvious in critical ill patients *(16-19)*. However, the relationship between lymphocyte reduction at the early stage and the severity and death risk of COVID-19 patients has not yet to be determined. Our results showed that the severity of COVID-19 patients was positively associated with lymphocyte reduction on admission. Moreover, fatality rate was higher in patients with lymphocyte reduction than those of patients with normal lymphocyte. These results indicate that lymphocyte reduction elevates the severity and death risk of COVID-19 patients, which explains why the fatality rate was higher in older COVID-19 patients than in younger ones. The present study provides evidence that lymphocyte reduction on admission may be a potential negative prognostic indicator for COVID-19 patients.

In summary, the present study analyzed the association between lymphocyte reduction at the early stage and prognosis of COVID-19 patients. We showed that older patients were more susceptible to lymphocyte reduction. Moreover, lymphocyte reduction on admission was positively associated with multiple organ injuries. In addition, lymphocyte reduction on admission elevated the severity and death risk of COVID-19 patients. Our results suggest that lymphocytes may be a direct target of SARS-COV-2. We provide evidence that lymphocyte reduction at early stage may be a potential negative prognostic indicator for COVID-19 patients. Therefore, surveillance of lymphocyte count is helpful for the early screening, diagnosis and treatment of critical ill COVID-19 patients.

## Data Availability

All data referred to in the manuscript are availability.

## Acknowledgments

We thank all patients and their families involved in this research. We also thank all members of respiratory and critical care medicine in the Second Affiliated Hospital of Anhui Medical University and Union Hospital of Huazhong University of Science and Technology for recruiting participators.

## Contributors

HZ, DXX, JF and LF designed research; LF, JF, HXX, YX, MDL, FFL and YL conducted research; LF analyzed data; HZ and JF wrote the paper; HZ and JF had primary responsibility for final content. All authors read and approved the final manuscript.

## Data and materials availability

All data associated with this study are present in the main paper.

## References

1. C. Huang, Y. Wang, X. Li, L. Ren, J. Zhao, Y. Hu, L. Zhang, G. Fan, J. Xu, X. Gu, Z. Cheng, T. Yu, J. Xia, Y. Wei, W. Wu, X. Xie, W. Yin, H. Li, M. Liu, Y. Xiao, H. Gao, L. Guo, J. Xie, G. Wang, R. Jiang, Z. Gao, Q. Jin, J. Wang, B. Cao, Clinical features of patients infected with 2019 novel coronavirus in Wuhan, China. Lancet 395, 497–506 (2020).

2. J. Wu, X. Wu, W. Zeng, D. Guo, Z. Fang, L. Chen, H. Huang, C. Li, Chest CT findings in patients with corona virus disease 2019 and its relationship with clinical features. Invest Radio doi: 10.1097/RLI.0000000000000670 (2020).

3. H. Shi, X. Han, N. Jiang, Y. Cao, O. Alwalid, J. Gu, Y. Fan, C. Zheng, Radiological findings from 81 patients with COVID-19 pneumonia in Wuhan, China: a descriptive study. Lancet Infect Dis pii: S1473–3099(20)30086-4 (2020).

4. N. Chen, M. Zhou, X. Dong, J. Qu, F. Gong, Y. Han, Y. Qiu, J. Wang, Y. Liu, Y. Wei, J. Xia, T. Yu, X. Zhang, L. Zhang, Epidemiological and clinical characteristics of 99 cases of 2019 novel coronavirus pneumonia in Wuhan, China: a descriptive study. Lancet 395, 507–513 (2020).

5. World Health Organization. https://www.who.int/emergencies/diseases/novel-coronavirus-2019 (2020).

6. D. Wang, B. Hu, C. Hu, F. Zhu, X. Liu, J. Zhang, B. Wang, H. Xiang, Z. Cheng, Y. Xiong, Y. Zhao, Y. Li, X. Wang, Z. Peng, Clinical Characteristics of 138 Hospitalized Patients With 2019 Novel Coronavirus-Infected Pneumonia in Wuhan, China. JAMA doi: 10.1001/jama.2020.1585 (2020).

7. J. J. Zhang, X. Dong, Y. Y. Cao, Y. D. Yuan, Y. B. Yang, Y. Q. Yan, C. A. Akdis, Y. D. Gao, Clinical characteristics of 140 patients infected with SARS-CoV-2 in Wuhan, China. Allergy doi: 10.1111/all.14238 (2020).

8. X. Xie, Z. Zhong, W. Zhao, C. Zheng, F. Wang, J. Liu, Chest CT for Typical 2019-nCoV Pneumonia: Relationship to Negative RT-PCR Testing. Radiology 200343 (2020).

9. M. Casarini, F. Ameglio, L. Alemanno, P. Zangrilli, P. Mattia, G. Paone, A. Bisetti, S. Giosuè, Cytokine levels correlate with a radiologic score in active pulmonary tuberculosis. Am J Respir Crit Care Med 159, 143–148 (1999).

10. W. J. Guan, Z. Y. Ni, Y. Hu, W. H. Liang, C. Q. Ou, J. X. He, L. Liu, H. Shan, C. L. Lei, D. S. C. Hui, B. Du, L. J. Li, G. Zeng, K. Y. Yuen, R. C. Chen, C. L. Tang, T. Wang, P. Y. Chen, J. Xiang, S.Y. Li, J. L. Wang, Z. J. Liang, Y. X. Peng, L. Wei, Y. Liu, Y. H. Hu, P. Peng, J. M. Wang, J. Y. Liu, Z. Chen, G. Li, Z. J. Zheng, S. Q. Qiu, J. Luo, C. J. Ye, S. Y. Zhu, N. S. Zhong, China Medical Treatment Expert Group for Covid-19, Clinical Characteristics of Coronavirus Disease 2019 in China. N Engl J Med doi: 10.1056/NEJMoa2002032 (2020).

11. C. Wu, X. Chen, Y. Cai, J. Xia, X. Zhou, S. Xu, H. Huang, L. Zhang, X. Zhou, C. Du, Y. Zhang, J. Song, S. Wang, Y. Chao, Z. Yang, J. Xu, X. Zhou, D. Chen, W. Xiong, L. Xu, F. Zhou, J. Jiang, C. Bai, J. Zheng, Y. Song, Risk Factors Associated With Acute Respiratory Distress Syndrome and Death in Patients With Coronavirus Disease 2019 Pneumonia in Wuhan, China. JAMA Intern Med doi: 10.1001/jamainternmed.2020.0994 (2020).

12. C. Qin, L. Zhou, Z. Hu, S. Zhang, S. Yang, Y. Tao, C. Xie, K. Ma, K. Shang, W. Wang, C. Xie, K. Ma, K. Shang, W. Wang, D. S. Tian, Dysregulation of immune response in patients with COVID-19 in Wuhan, China. Clin Infect Dis pii: ciaa248 (2020).

13. K. Liu, Y. Y. Fang, Y. Deng, W. Liu, M. F. Wang, J. P. Ma, W. Xiao, Y. N. Wang, M. H. Zhong, C. H. Li, G. C. Li, H. G. Liu, Clinical characteristics of novel coronavirus cases in tertiary hospitals in Hubei Province. Chin Med J doi: 10.1097/CM9.0000000000000744 (2020).

14. F. Song, N. Shi, F. Shan, Z. Zhang, J. Shen, H. Lu, Y. Ling, Y. Jiang, Y. Shi, Emerging Coronavirus 2019-nCoV Pneumonia. Radiology 200274 (2020).

15. Y. Liu, Y. Yang, C. Zhang, F. Huang, F. Wang, J. Yuan, Z. Wang, J. Li, J. Li, C. Feng, Z. Zhang, L. Wang, L. Peng, L. Chen, Y. Qin, D. Zhao, S. Tan, L. Yin, J. Xu, C. Zhou, C. Jiang, L. Liu, Clinical and biochemical indexes from 2019-nCoV infected patients linked to viral loads and lung injury. Sci China Life Sci 63 364–374 (2020).

16. P. Zhou, X. L. Yang, X. G. Wang, B. Hu, L. Zhang, W. Zhang, H. R. Si, Y. Zhu, B. Li, C. L. Huang, H. D. Chen, J. Chen, Y. Luo, H. Guo, R. D. Jiang, M. Q. Liu, Y. Chen, X. R. Shen, X. Wang, X. S. Zheng, K. Zhao, Q. J. Chen, F. Deng, L. L. Liu, B. Yan, F. X. Zhan, Y. Y. Wang, G. F. Xiao, Z. L. Shi, A pneumonia outbreak associated with a new coronavirus of probable bat origin. Nature 579: 270–273 (2020).

17. R. Qu, Y. Ling, Y. H. Zhang, L. Y. Wei, X. Chen, X. Li, X. Y. Liu, H. M. Liu, Z. Guo, H. Ren, Q. Wang, Platelet-to-lymphocyte ratio is associated with prognosis in patients with Corona Virus Disease-19. J Med Virol doi: 10.1002/jmv.25767 (2020).

18. R. Han, L. Huang, H. Jiang, J. Dong, H. Peng, D. Zhang, Early Clinical and CT Manifestations of Coronavirus Disease 2019 (COVID-19) Pneumonia. AJR Am J Roentgenol 1–6 (2020).

19. Liu F., A. Xu, Y. Zhang, W. Xuan, T. Yan, K. Pan, W. Yu, J. Zhang, Patients of COVID-19 may benefit from sustained lopinavir-combined regimen and the increase of eosinophil may predict the outcome of COVID-19 progression. Int J Infect Dis pii: S1201–9712(20)30132-6 (2020).

